# Understanding staff needs for Improving End-of-life Care in Critical Care Units

**DOI:** 10.1101/2024.02.09.24302454

**Authors:** S Tavabie, S Pearson, J Balabanovic, A Batho, M Juj, P Kastande, J Bennetts, E Collis, T. Bonnici

## Abstract

**Objectives:** Critical care is a place of frequent death, up to a quarter of those admitted die during admission. Caring for dying people provides many challenges, practically, professionally and personally. The aim of this study was to better understand the perspectives of staff caring for dying people in critical care and identify their priorities for improvement.

**Method:** Three multidisciplinary focus groups of critical care staff at a large central London hospitals trust were facilitated with a semi structured format and digitally transcribed. Inductive thematic analysis was conducted to extract themes.

**Results:** N=34 (18 nursing, 7 allied health professionals, 6 medical, 3 clerical/administrative) The five themes were structured as priority statements: “We need to recognise” included the subthemes of being “sick enough to die” and potential rapid deteriorations in this setting; “We need to understand” with subthemes of perspectives on dying and prioritising time for conversations; “We need to connect” with subthemes of therapeutic relationship and physical presence; “We need to collaborate” with subthemes of critical care working and empowerment, and cross teams working; “We need support” with themes of experiencing support and making time to support others.

**Conclusion:** We present an approach to identifying critical care departmental priorities for an end-of-life care improvement programme. The themes extracted will be used to evaluate systems for dying in critical care, aiming to empower staff to provide excellent care every time they look after a dying person.

## Introduction

*“Her phone was still on, but it was on vibrate and somebody had put it in the drawer. Her mum kept ringing” (P5 Nurse)*

The concept of a ‘good death’ is multifaceted, using domains within the holistic model of care developed by specialists in palliative care (1). However, current discourse outlines the inadequacies of the ‘good death’ narrative instead focusing on the qualities of end-of-life experience or ‘matters of care’ (2). When considered in this manner, the quality of dying and the care one receives can be evaluated from the perspectives of those involved within an end-of-life system (3). When considering those persons admitted to the critical care unit, the primary stakeholders in a death can broadly be grouped into the dying person, those important to them, their communities, and those staff working in the Intensive Care Unit (ICU), from the bedside nurse, the leading consultant, the administrative staff, allied health professionals and many others.

Critical care is a place of frequent death with 15-25% of people admitted to ICU dying during their stay, amounting to 10-20% of the population of the UK (4). The trajectory of death in the ICU is often unpredictable and influenced by various factors, including severity of illness; presence of comorbidities, and effectiveness of treatment. In some cases, patients may have a sudden and unexpected decline in their health status, whereas in others, the decline may be gradual over a longer period (5). The trajectory of death can also be affected by the decisions made by patients, families, and healthcare providers with approximately 70% of deaths following the withdrawal or withholding of life-sustaining treatment (6; 7).

Caring for dying people can be one of the most stress provoking experiences for healthcare professionals (8). Being unable to provide care that is congruous with your own moral values leads to increased risk of moral injury (9). Particularly during the COVID-19 pandemic, staff across all sectors of health and care supported patients in challenging circumstances and left them with distressing memories (9). The focus of different staff groups when considering high quality care for dying people in an ICU environment differ, with medical staff often focusing on pain and symptom management and clear communication, and nursing staff focusing on psychosocial and spiritual support, although both groups acknowledge the importance of each other’s roles (10) (11) (12). There is little to describe the priorities of all staff within a unit, nor is there an accepted approach to discover these.

This project aimed to understand the perspectives and priorities of staff of all backgrounds working in the critical care units of a large, central London, acute and specialist services NHS Foundation Trust.

## Method

### Aim

To explore the experiences and priorities of staff caring for dying people in critical care.

### Setting

The NHS provides free at the point of delivery healthcare for all (13). Admission to ICU for life-sustaining supportive care during critical illness is determined by the managing clinicians after appraisal of needs, likelihood of morbidity and mortality and patient priorities (14). The ICU is a naturally multidisciplinary environment with a broad range of disciplines working together to provide care holistically. University College London Hospital’s (UCLH) adult general and haemato-oncology ICUs provide care to ∼1700 patients annually. UCLH is situated in the central London borough of Camden, a diverse population with over 140 registered languages and dialects.

### Design

A multidisciplinary end-of-life care in critical care improvement group with representatives from senior and junior critical care medical, nursing, AHP, and support staff, as well as specialist palliative and end-of-life care, chaplaincy and patient representation was convened. A semi-structured focus group script was developed following literature review. Agreement on transcription, shared facilitation and faculty reflexivity made (Appendix 1 – Focus group script). Transcription was performed automatically using the Microsoft Teams™ function and anonymised. One faculty member also provided supplementary annotation to ensure capture of emotion in the discussion.

### Participation

All staff working in general and haemato-oncology ICUs were invited using email, poster, and direct approach through handover meetings. Inclusion criteria were simply of current employment in ICU and willingness to participate. There were no exclusion criteria. To ensure open contribution and reduce barriers to participation, only staff group was collected for demography. 3 focus groups were held at varying times in the day, and different days in the week, over a 2-month period in February-March 2023

### Reflexivity

Reflexivity is vital within qualitative research and involves the process of “reflecting critically on oneself as a researcher” (15). A reflective log was maintained by the project lead and faculty were invited to reflect together after each focus group and at fortnightly meetings for the duration of the project.

### Data analysis

The focus group transcripts were evaluated using an inductive approach using a reflexive thematic analysis as described by Braun and Clark, and themes were extracted from the data (16). Analysis was undertaken manually by the project lead, codes were distributed into a coding tree of themes and subthemes, and refined and reviewed at regular intervals. Further validity was gained through the review of themes with project supervisors.

## Results

34 participants with wide variations in seniority and professional background (18 NURSING, 7 allied health professionals, 6 medical, 3 clerical/administrative). Figure 1 provides a visual overview of the themes formed following inductive thematic analysis of transcripts from the focus groups.

**Figure 1:**
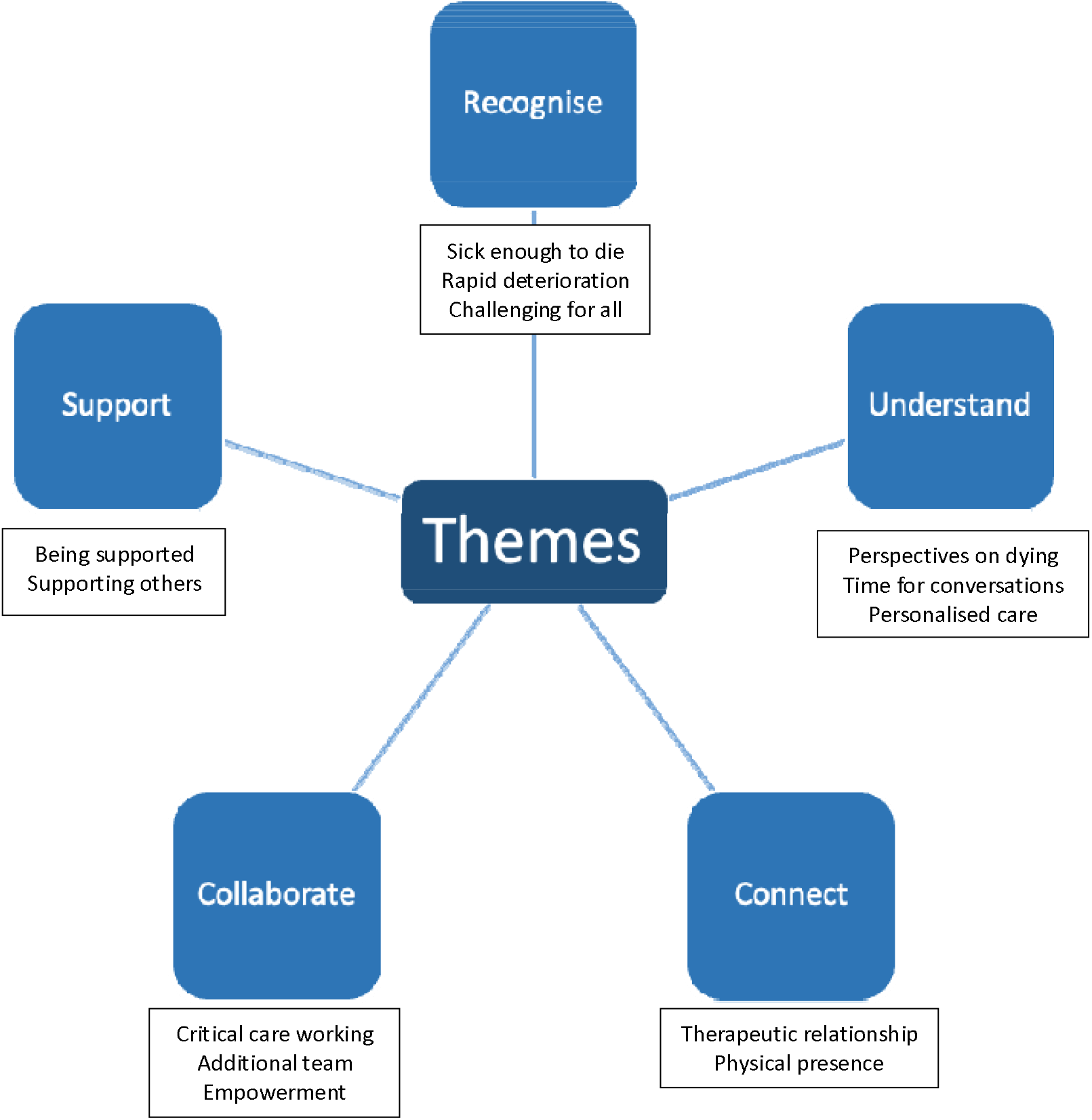
Visual representation of themes extracted from focus group data

### Theme 1 “Recognise”

#### Subtheme: “Sick enough to die”

Patients requiring critical care are more often than not, “sick enough to die.” Differentiation between those patients who will go on to die, and those who will survive is therefore very challenging. Some participants reflected on experiences where care was better because of earlier clarity of outcome, while others felt sharing a recognition of this difficulty aided earlier conversations and preparations.

*“It was determined very early on between the primary team and ITU that this person wasn’t going to survive this insult so we got palliative care involved very, very early on and…she was very peaceful, very comfortable and it was one of those deaths that the preplanning worked very well” P5 Nurse*

*“If we understand at some point that this (death) might be a possibility…we [should] sit down with the family and say “things at the moment are not doing well, there’s a possibility that things continue to get worse and with that perspective, what is it that you would want… is there something we can do to support you or something you would want us to know early on?” P30 Doctor*

#### Subtheme: “Rapid deterioration”

Participants often reflected that, in their experience, people in the intensive care unit did not give the ‘warning signs’ that ward patients might do, causing more challenges in recognition of dying. Escalation planning was commented on as a frequent practice, but participants reflected in their perceived lack of sharing that, were the patient to deteriorate beyond the appropriate level of support, this would mean they were dying.

*“…he just got so sick so quickly…we knew things were getting worse, but one-minute things were ok, then suddenly everything’s flashing and beeping and well…a few minutes later he had died…’ P22 Nurse*

*“I think what gets hard for us is when we’re still kind of actively treating but not fully comfort care*… *Can you treat as long as the patient’s comfortable? Don’t do anything aggressive” etc… it’s hard for everyone but it’s kind of blurred instructions for us” P27 Physiotherapist*

#### Subtheme: “Challenging for all”

It was recognised that where senior clinicians made decisions based on their experience and expertise, it was challenging for every member of the team. Deaths during the COVID-19 pandemic were described as particularly difficult to predict because there was no experience in managing these patients before their deterioration.

*“We get pulled into doing interventions where actually, in hindsight, 2 days down the line when the patient dies, it becomes very easy to say we shouldn’t have done that. But at that moment when you’re making a decision to…you know there’s maybe a 1% chance that maybe the patient can survive” P10 Doctor*

*“a good death on ITU is a really difficult thing, cause we’re constantly…pulled between these two polar opposites which often come across as binary treatment streams where you do everything from filter and invasively ventilate to do nothing” P30 Doctor*

### Theme 2: “Understand”

#### Subtheme: “Perspectives on dying”

Staff reflected that many patients and their relatives have different perspectives on death and dying, where and how death might best occur and the level of medical intervention appropriate to try to prevent it. Understanding this often allowed them to care for people and provide information in a manner congruous with what was medically appropriate and sensitive to their individual priorities.

*“They weren’t ready to hear it for ages but then towards the end they could hear it. I think we’d done the groundwork and so at the time that he died they were prepared, and they knew*.*” P11 Nurse*

*“Sometimes it’s so straight forward and the family get you right away, they know that their loved one is dying. Other times they want you to keep going on and on and on and you have to try and help them understand that’s not okay” P26 Nurse*

#### Subtheme: “Time for conversations”

Participants reflected on the barriers to developing an understanding of the person and their loved ones’ perspectives and priorities around dying, notably time and space. They felt that where these were overcome, often at personal cost to the staff member, a better understanding was reached, and care was better overall.

*“Conversations had over the phone, or even in a corridor feel wrong. When you get to take the time and do it properly, you can see the difference it makes for (the family)” P10 Doctor*

“(When it’s busy) *it’s so easy to walk past those side rooms without even going in to see what’s the symptom control like and meet the family to really put a face to the name… you just walk past the room and say end-of-life pathway and move on*.*” P30 Doctor*

#### Subtheme: “Personalised care”

It was discussed that a personalised, holistic care plan was helpful but often challenging to formulate in a stressful environment. Situations often demanded greater flexibility and pragmatism, focusing on the possible rather than the ideal.

*“having it [individualised care plan for the dying person] is useful but normally it’s all too quick and so we focus on what we can do then and there, rather than looking at forms or whatever” P26 Nurse*

### Theme 3: Connect

#### Subtheme: Therapeutic relationship

HCPs felt that there was a significant benefit to connecting with the ‘premorbid person’ and their loved ones. Connecting with their life before hospital allowed them to better empathise and provide personalised care and allowed them to feel professionally fulfilled. Conversely, where care happened without this, such as during the peaks of the COVID-19 pandemic, care was felt to suffer.

*“We are connected to the patient, connected to the family, and really make a difference, but that connection wasn’t there [in the pandemic]. The absence of that made it really hard because, that’s not what we do in critical care*… *When you think of those terrible, terrible deaths…it’s the connection that wasn’t there” P8 Nurse*

#### Subtheme: Physical presence

Staff felt that the COVID-19 pandemic had added extra weight to the significance of having someone in the room with the dying person. Nursing staff in particular frequently felt that having someone in the room, holding the person’s hand was extremely important to ensure a ‘good death.’ Where families were unable to be there because of travel difficulties, this was perceived as a particular stress point.

*“The nurse actually rang me during the day and told me she’d gone peacefully. She sat there, holding her hand [as I had asked her to]. For me, that will always be there. I’m never ever gonna forget…” P7 Nurse*

*“the curtains were around, she was dying, and no one was there. She died on her own. I always remember how awful we felt because we were so busy. No one was with her. I always think it’s really important to just be there to hold their hands” P13 Nurse*

### Theme 4: Collaborate

#### Subtheme: Critical Care Working

It was often expressed that the nature of critical care was as an environment in which specialist support to organs allowed for the management of conditions often guided by other specialty teams. As such, the difficulty in balancing the priorities of the critical care and ‘primary team’ was discussed.

*“As long as the primary team wants to drive things forward, I mean, who are we to say this is not to continue…” P9 Doctor*

#### Subtheme: Additional teams

Participants explored the value of ‘additive’ teams such as those of specialist palliative and end-of-life care and chaplaincy.

*“the priest came in and we all organised together with the boyfriend that there was some sort of…celebration of them, the love they have for each other and so on…she didn’t look at all like a patient, just her normal self*…*She was happy, the family was happy, everyone was…was celebrative…” P13 Nurse*

#### Subtheme: Empowerment

In each of the focus groups, participants raised the importance of each member of the multidisciplinary team feeling able to raise concerns and to be nurtured positively in their contributions to care. It was frequently raised that ward nurses were not included in breaking bad news conversations, and this made it harder for them to speak up and to care for the dying person.

*“I feel I didn’t provide the support to our team on the ground on that shift to say, ‘it’s okay,’ you know. They didn’t feel empowered to be able to speak up and say, is this the right thing to do? Should we be doing this?” P30 Doctor*

*“I don’t feel powerful enough to have [conversations about dying]. I feel that sometimes, perhaps, even if I…share that I think this patient might be dying, it’s not recognised from the medical team…If I share my opinion, maybe [this would help], but I wish it would be easier to get to that point” P18 Nurse*

*“[If I had been included in that meeting] I would have been able to put my point across and make them understand. I would have been able to have a clear plan in my head” P13 Nurse*

### Theme 5: Support

#### Subtheme: Being supported

Staff reflected that support for the individuals’ providing care was often lacking. Structures to allow staff to take time to decompress, debrief, reflect and learn were lacking, particularly during times of stress to the service.

*“the absolutely brutal deaths of COVID, I don’t know how many dead patients I saw during the pandemic…we couldn’t support each other, we couldn’t support their families…I could see people getting immune and it broke my heart…you go into self-protection” P8 Nurse*

#### Subtheme: Supporting others

It was felt that there was a specific benefit in offering emotional, psychological, and practical support to colleagues dealing with difficulties following caring for a dying person. There were several occasions where this was offered during the focus groups, and when reflecting, participants revealed that this was helpful for their own processing, but that there were significant barriers to doing so while at work.

*“I could hear in your voice that you’re carrying this young girl with you*…*you were hurting for her for not having her mum there, and for her mother too, you’re a mother too…you were there*.*” P19 Nurse*

*“Yeah, I really feel it when I see someone going through it, I want to reach out and be there for them. It hurts to see someone suffering like that” P26 Nurse*

## Discussion

In this focus group study, we explored the perspectives and priorities for improvement of staff caring for dying people in critical care settings in a central London acute and specialist services NHS Foundation Trust. Through inductive thematic analysis we have identified 5 themes outlining the priorities of critical care staff, presented with subthemes through the previous section. As agreed, these themes are expressed as ‘needs’ to drive improvement in the critical care units. “We need to recognise,” “We need to understand,” “We need to connect,” “We need to collaborate,” “We need support.”

Lack of recognition has been shown to be a key barrier to high quality end-of-life care and despite significant work to better prognosticate, it continues to present an issue (17). Educational interventions have been shown to improve the ability of staff to recognise dying and given UCLH’s ‘Transforming End of Life Care’ and Critical Care education teams, this has been a first focus for improvement (18).

In our focus groups, nurses have expressed concern about being excluded from discussions with families about bad news, which are usually led by the ICU consultant and not at the bedside. This situation increases the nurses’ anxiety about what they can communicate to a family and a feeling of withholding information, either because they are not aware of the medical plan or unsure about what has been shared with the family (19). Nurses often feel underprepared for critical conversations about end of life care and though common practice is for role-play exercises, the complexity of conversations in critical care demands a combination of experience, empathy and reflective practice (8) (20) (21). The COVID-19 pandemic may have lessened the inclusion of nursing staff in these critical conversations due to ‘social distancing’ and staffing issues. Our findings have encouraged us to move back to pre-pandemic practice of inclusion of nursing staff, enhancing their ability to connect with those in their care, advocate on their behalf, learn through reflective participation and provide individualised end-of-life care.

Understanding individual perspectives, cultural and social expectations, and personal history of those in our care allows us to facilitate truly personalised care, beyond the person’s biology; however, the challenges of doing so, particularly in a critical care setting, are well described (22). The patient – carer relationship has been the subject of extensive study, with significant benefit felt when a connection can be nurtured, and detriment when it is missing (23). The end-of-life care quality improvement group functions as an integral component of the larger Critical Care Patient Experience group at UCLH. This arrangement facilitates a collaborative approach to monitoring the experiences of patients and their families in the critical care setting. Furthermore, it ensures that improvement initiatives are thoughtfully aligned with the principles of personalised care, reflecting a comprehensive approach to enhancing patient and family experiences in critical care.

Many critical care units in the United Kingdom operate a ‘closed unit’ philosophy, where the critical care team takes ultimate responsibility for management of the patient and therefore, of significant decision making around their care, supported by ‘primary’ and ‘secondary’ specialist teams - giving weight to staff’s calls for more effective collaboration (24). Specialist palliative and end-of-life care teams can provide support to critical care in consultative (as in UCLH) or and integrated capacity (25). The priorities presented in this project are those of staff in critical care, not in aforementioned ‘primary’ or ‘secondary’ teams, nor those of the loved ones of those who die in critical care, nor of the patients themselves. Staff perspectives are shaped by their caregiving experiences and therefore provide anecdotes, alongside a general overview of care delivered. They sit alongside the 5 priorities of the dying person (recognise, communicate, involve, support, plan and do) developed by the Leadership Alliance for the Dying Person when considering (26). Our final theme of “Support” is unsurprising given our focus on staff perspectives and priorities. This represents a significant addition when considering our themes as a guide for quality improvement, in that offering support to staff and space that they might support each other is felt to be key to ensuring high quality end-of-life care. Previous research has investigated specific aspects of a death that may be more or less associated with the perception of ‘good deaths’ in critical care, however our framing as ‘staff needs’ to guide improvement is novel (10).

### Limitations

This is a small, focus group-based study in one organisation, operating in a publicly funded, free at the point of delivery, critical care department, serving highly specialist and acute care in the centre of a capital city. As such, our findings have limited generalisability. However, they do represent the views expressed within the groups themselves, and through the open invitation, limited barriers to participation and facilitated, semi-structured design of the sessions, generalise to our critical care units more broadly.

Within the limitations, it is important to be aware of the researcher’s role within the research itself, which may introduce an element of bias. As the project lead was a palliative medicine doctor working within the transforming end-of-life care team, one could argue that this may have added an element of bias to the project design, although this was mitigated by the multidisciplinary quality improvement group and facilitation of sessions.

## Conclusion

We have presented an approach that allowed for the extraction of 5 themes, framed as the priorities of staff for quality improvement in the care of dying people in our critical care units. Further work should be conducted to explore differences across other units, considering patient and staff demography, service makeup and skill mix. Though conversations with those that sadly die are impossible, these staff priorities could be explored with the bereaved, or those discharged from critical care to investigate whether they would be congruous with their own. The themes extracted from our focus groups each allow for focused innovation and service development to facilitate excellent end-of-life care in our critical care units, even in the face of adversity.

## Data Availability

All data produced in the present study are available upon reasonable request to the authors

## Governance and ethics

As per HRA Research decision tool this study was conducted as a part of service improvement, does not report generalisable findings nor directly involve patients and as such does not require ethical approval.

As a quality and service improvement project it was registered through Critical Care clinical governance at UCLH

## Declarations

ST, SP, EC and TB had the original idea for the project. All authors contributed to the design of the focus group scripts and/or acted as faculty for the groups themselves. ST embedded in and coded the data, performing analysis. ST and SP prepared the initial manuscript with all authors contributing to this prior to submission.

## Thanks

We would like to express our thanks for the significant contributions made to the project by: Sadaf Iqbal, Katie Whelan, Dr Kate Urwin, Dr Suzanne Benhalim, Dr Zoe Burgess, and Dr Jon Donaghy.

We would also like to share our thanks to all the staff working hard, caring for people who are “sick enough to die” across critical care units at UCLH, particularly those who took part in our focus groups without whom this study could not have taken place.

## Notes

### Competing Interest Statement

The authors have declared no competing interest.

### Funding Statement

This study did not receive any funding

### Author Declarations

In conducting our study, we recognised the paramount importance of ethical oversight. The ethical review for our project was undertaken by the University College London Hospitals NHS Trust (UCLH) Critical Care Multidisciplinary Integrated Governance (MIG) meeting. UCLH is a renowned health institution located in England, UK. The UCLH Critical Care MIG carefully reviewed our study and concluded that formal ethical approval was not necessary. This decision was based on the classification of our project as service evaluation, aligning with the 2023 guidelines provided by the Health Research Authority (HRA) in England. According to these guidelines, our project did not fall under the category of 'research', thereby not requiring HRA approval. Despite this waiver, we adhered rigorously to the United Kingdom Policy Framework for Health and Social Care Research, as outlined by the HRA in 2023. This adherence underscores our commitment to maintaining ethical standards in conducting evaluations, ensuring the safeguarding of staff and participants involved in our study. Additionally, it is important to note the process of participant engagement in our project. We provided clear information regarding the purpose of the engagement events. Participation in these events was based on implicit consent, with the understanding that attendees were fully informed and their engagement constituted consent to participate. In summary, while our study was exempt from formal ethical approval by the UCLH Critical Care MIG, due to its classification as service evaluation, we took all necessary steps to ensure ethical integrity. Our adherence to the UK's established health and social care research policies reflects our unwavering commitment to ethical research practices.

